# Gestational SARS-CoV-2 infection is associated with placental expression of immune and trophoblast genes

**DOI:** 10.1101/2022.02.22.22271359

**Authors:** Corina Lesseur, Rebecca H. Jessel, Sophie Ohrn, Yula Ma, Qian Li, Fumiko Dekio, Rachel I. Brody, James G. Wetmur, Frederieke A.J. Gigase, Molly Lieber, Whitney Lieb, Jezelle Lynch, Omara Afzal, Erona Ibroci, Anna-Sophie Rommel, Teresa Janevic, Joanne Stone, Elizabeth A. Howell, Romeo R. Galang, Siobhan M. Dolan, Veerle Bergink, Lotje D. De Witte, Jia Chen

**Author notes:** **Corresponding Author:** Jia Chen, ScD. Department of Environmental Medicine and Public Heath, Icahn School of Medicine at Mount Sinai, 1 Gustave L. Levy Place, Box 1057, New York, NY 10029, USA.

## Abstract

**Introduction:** Maternal SARS-CoV-2 infection during pregnancy is associated with adverse pregnancy outcomes and can have effects on the placenta, even in the absence of severe disease or vertical transmission to the fetus. This study aimed to evaluate histopathologic and molecular effects in the placenta after SARS-CoV-2 infection during pregnancy.

**Methods:** We performed a study of 45 pregnant participants from the Generation C prospective cohort study at the Mount Sinai Health System in New York City. We compared histologic features and the expression of 48 immune and trophoblast genes in placentas delivered from 15 SARS-CoV-2 IgG antibody positive and 30 IgG SARS-CoV-2 antibody negative mothers. Statistical analyses were performed using Fisher’s exact tests, Spearman correlations and linear regression models.

**Results:** The median gestational age at the time of SARS-CoV-2 IgG serology test was 35 weeks. Two of the IgG positive participants also had a positive RT-PCR nasal swab at delivery. 82.2% of the infants were delivered at term (≥37 weeks), and gestational age at delivery did not differ between the SARS-CoV-2 antibody positive and negative groups. No significant differences were detected between the groups in placental histopathology features. Differential expression analyses revealed decreased expression of two trophoblast genes (*PSG3* and *CGB3*) and increased expression of three immune genes (*CXCL10, TLR3* and *DDX58*) in placentas delivered from SARS-CoV-2 IgG positive participants.

**Discussion:** SARS-CoV-2 infection during pregnancy is associated with gene expression changes of immune and trophoblast genes in the placenta at birth which could potentially contribute to long-term health effects in the offspring.

## 1. Introduction

The current pandemic of coronavirus disease 2019 (Covid-19) is caused by infection with the novel severe acute respiratory syndrome coronavirus 2 (SARS-CoV-2). As of January 24, 2022 approximately 352 million infections and 5.6 million deaths have been reported worldwide [1]. The clinical manifestations of SARS-CoV-2 infection vary widely by age group and presence of comorbid conditions, ranging from asymptomatic infection to respiratory failure, multisystem organ failure, and death in critically ill patients [2, 3]. Although comprehensive knowledge of the factors involved in severe Covid-19 is still needed, dysregulation of the host inflammatory and immune responses [4, 5] and thrombosis [6] have been implicated in Covid-19 induced tissue damage.

During pregnancy, the maternal immune system undergoes a series of dynamic changes aimed to promote tolerance of the fetus, that can also influence responses to pathogens, including viruses [7]. Recent reports show that pregnant individuals with severe SARS-CoV-2 infection are at higher risk of intensive care unit admission, mechanical ventilation, extracorporeal membrane oxygenation, and mortality compared to non-pregnant individuals [8, 9]. Other studies show that active SARS-CoV-2 infection at delivery (mainly confirmed through a PCR positive test) is associated with obstetric and neonatal complications including increased risk of preterm birth, stillbirth, miscarriage, preeclampsia, emergency cesarean section and higher neonatal morbidity [8, 10-18]. However, in other reports, including ours from New York City and a Denmark study, SARS-CoV-2 IgG seropositivity without RT-PCR positivity at delivery was not associated with adverse pregnancy outcomes [19, 20].

The main cell entry pathway of the SARS-CoV-2 virus is dependent on the *ACE2* receptor and aided by the *TMPRSS2* proteases of the host cells [21]. Early in pregnancy, *ACE2* and *TMPRSS2* are expressed in placental cytotrophoblast and syncytiotrophoblast cells; however, the expression of these proteins is low in term placentas [22-24]. Recent data suggest that multiple placental cell types are susceptible to infection in explants and immortalized cells after exposure to SARS-CoV-2, and infection susceptibility is related to the levels of *ACE2* expression [25-27]. Yet, existing data show that vertical transmission of SARS-CoV-2 is rare; only a few reports have documented presence of the virus in the fetal compartments of the placenta or in newborns [28-30]. Moreover, most of the histopathology studies suggest that placental infection with SARS-CoV-2 is also rare [31-33] and can exist in the absence of vertical transmission [34, 35]. Additionally, the criteria to diagnose SARS-CoV-2 placental infection have been inconsistent across studies [34, 36-40]. Nonspecific placental histopathologic lesions have also been associated with maternal SARS-CoV-2 infection; a recent meta-analysis reports increases in risk of fetal vascular malperfusion, acute and chronic proinflammatory lesions, increased perivillous fibrin, and intervillous thrombosis [41]. These lesions could result from localized placental SARS-CoV-2 infection and/or inflammatory responses to the systemic maternal infection. However, most of the available reports evaluated placental effects of acute SARS-CoV-2 infections mainly from mothers infected at delivery (positive nasopharyngeal PCR test). To date, only few studies have reported changes in expression of immune and inflammatory genes in the placenta, also in cases of acute SARS-CoV-2 infection [42-44]. Understanding the impact of maternal SARS CoV-2 infection on the placenta during pregnancy, including among participants without active infection at delivery, is vital because these placental changes can lead to adverse pregnancy outcomes and long-term effects on the health of newborns [45]. The aim of this work was to evaluate histopathologic and gene expression changes in placentas delivered from SARS-CoV-2 IgG positive compared to those from SARS-CoV-2 IgG negative pregnant individuals.

## 2. Methods

### 2.1 Study Population

The Generation C study is a prospective pregnancy cohort study that aims to examine the impact of SARS-CoV-2 infection during pregnancy on obstetric and neonatal outcomes. Pregnant individuals were recruited at Mount Sinai Hospital (MSH) and Mount Sinai West (MSW) in New York City (NYC) starting April 20, 2020, and recruitment is ongoing [19]. The first COVID-19 case in NYC was officially confirmed on March 1, 2020. Maternal blood samples are collected at multiple time points as part of routine clinical care. Electronic medical record (EMR) review and serological SARS-CoV-2 IgG tests are used to confirm past SARS-CoV-2 infection. Serological testing for IgG antibodies against the SARS-CoV-2 spike (S) protein (anti-S IgG) was performed using an enzyme-linked immunosorbent assay (ELISA) developed at the Icahn School of Medicine at Mount Sinai [46]. Placental samples are collected after delivery by the Mount Sinai Biorepository and Pathology Core. All participants provided written informed consent per the institutional review board (IRB)-approved study protocol (IRB at the Icahn School of Medicine at Mount Sinai, protocol IRB-20-03352, April 15, 2020).

For this analysis, we examined a subset of Generation C participants with and without evidence of past SARS-CoV-2 infection with available placenta tissue blocks collected for medical pathology examination and consent to donate placental tissue; 15 participants were SARS-CoV-2 IgG positive and 30 were SARS-CoV-2 IgG negative. EMR review was conducted to obtain clinical and sociodemographic characteristics of mothers and infants. Participants in this analysis gave birth between May and September of 2020. Since widespread community transmission of SARS-CoV-2 in NYC began in March 2020, we theorized that IgG positive participants who gave birth before or in late September were infected at some point during pregnancy. Additionally, all 45 pregnant participants delivered before the first COVID-19 vaccine received Emergency Use Authorization by the U.S. FDA in December 2020.

### 2.2 Placenta Histopathology

After fixation, placentas were processed according to standard protocols including comprehensive gross tissue and histopathologic examination of the umbilical cord, chorionic membranes, and placental villi. Histopathologic review was performed according to the Amsterdam Placental Workshop Group Consensus Statement guidelines [47]. All placentas in the study were reviewed for medical pathology and findings were recorded in the pathology report and in the EMR.

### 2.3 Targeted placental gene expression profiling

RNA was extracted from formaldehyde-fixed paraffin embedded (FFPE) tissue blocks using the Maxwell® 16 LEV RNA FFPE Purification Kit (Promega, Madison, WI). RNA concentration was determined using the Nanodrop (Thermo Fisher Scientific, MA). Gene expression was profiled with a custom designed NanoString codeset (NanoString, Seattle, WA) panel with 50 probes including genes involved in the inflammatory and/or immune response (n=25), stress response (n=8), cell-type markers (n=7), SARS-CoV-2 host response (n=6), viral SARS-CoV-2 genes (nucleocapsid and envelope proteins) and two housekeeping genes (*RPL19, RPLP0*) (**Supplementary Table 1**). RNA (100 nanograms) was hybridized overnight to reporter and capture probes at 65°C. Next, unbound probes were removed, and purified complexes were aligned and immobilized on four NanoString cartridges using the nCounter Prep station. Cartridges were scanned for gene counts detection in the nCounter Digital Analyzer. All laboratory protocols we performed following manufacturer’s instructions. Raw gene expression counts were imported from RCC files using the NanoStringNorm R package (1.2.1.1) [48]. To normalize CodeCount technical variation we used the geometric mean. Background expression levels were calculated based on the mean +/- 2SD of negative control probes. Values below the background limit of detection (LOD) for each sample (mean +/- 2SD of negative control probes) were replaced with LOD/√2. We used the geometric mean of the housekeeping genes to normalize for sample RNA sample content. After normalization, counts were log2 transformed for statistical analyses. Samples with less than 50% of probes above background were excluded (n=1, SARS-CoV-2 IgG negative), and probes with counts below the background level in more than 50% of the samples in each study group were removed (n=9). We used the placental-cell gene markers in the panel to calculate cell-type scores as the average of the log2 normalized expression of each of the cell type gene marker (*PEG10* and *PEG3* for cytotrophoblasts, *CGB3* and *PSG3* for syncytiotrophoblasts, *CD68* and *CD163* for macrophages and *PECAM1* for endothelial cells).

### 2.4 Statistical analyses

We used summary statistics including median, range, or frequency tables to evaluate the distribution of continuous and categorical variables. We performed bivariate analyses using Fisher exact tests and Wilcoxon signed-rank or Kruskal–Wallis tests as appropriate to evaluate differences in clinical, sociodemographic, and histopathology variables between the SARS-CoV-2 IgG positive versus negative groups. We used principal components analyses to evaluate possible effects of technical (e.g., NanoString cartridge) or biological (e.g., infant sex) covariates in placental gene expression. Differential gene expression analysis by SARS-CoV-2 IgG status was performed using the Limma R package [49] that uses an empirical Bayes method to fit linear models with moderated standard errors for each gene (continuous outcome variable) and the study group (IgG positive versus negative) as the predictor variable. We considered as possible confounders covariates that could influence placenta gene expression including infant sex, gestational age at birth, maternal age, and gestational age at SARS-CoV-2 IgG antibody test. We fitted linear models adjusted for covariates and cell-type proxy scores. Sensitivity analyses were performed excluding two participants with acute infections at delivery to evaluate the impact of SARS-CoV-2 PCR positivity at delivery. Statistical significance was set at *p*≤0.05. Analyses were performed in R statistical computing software version 4.1.0 [50].

## 3. Results

### 3.1 Demographic, clinical and placenta histopathologic characteristics

**Table 1** displays the characteristics of the Generation C participants included in these analyses (n=45) stratified by study groups: SARS-CoV-2 IgG negative (n=30) and SARS-CoV-2 IgG positive (n=15). The median gestational age of IgG serology testing was 35 weeks with an interquartile range (IQR) between 17.4 and 40.9 weeks. Two of the IgG positive participants were also SARS-CoV-2 PCR positive (nasopharyngeal swab) at the time of the labor and delivery admission. Like in the larger Generation C cohort [19], IgG seropositive participants had higher pre-pregnancy BMI (*p*=0.05) and were more often Hispanic, or non-Hispanic Black compared to IgG seronegative participants. In contrast, we did not observe differences in other maternal characteristics including age, parity, tobacco use, medical conditions, or obstetric conditions.

**Table 1.**
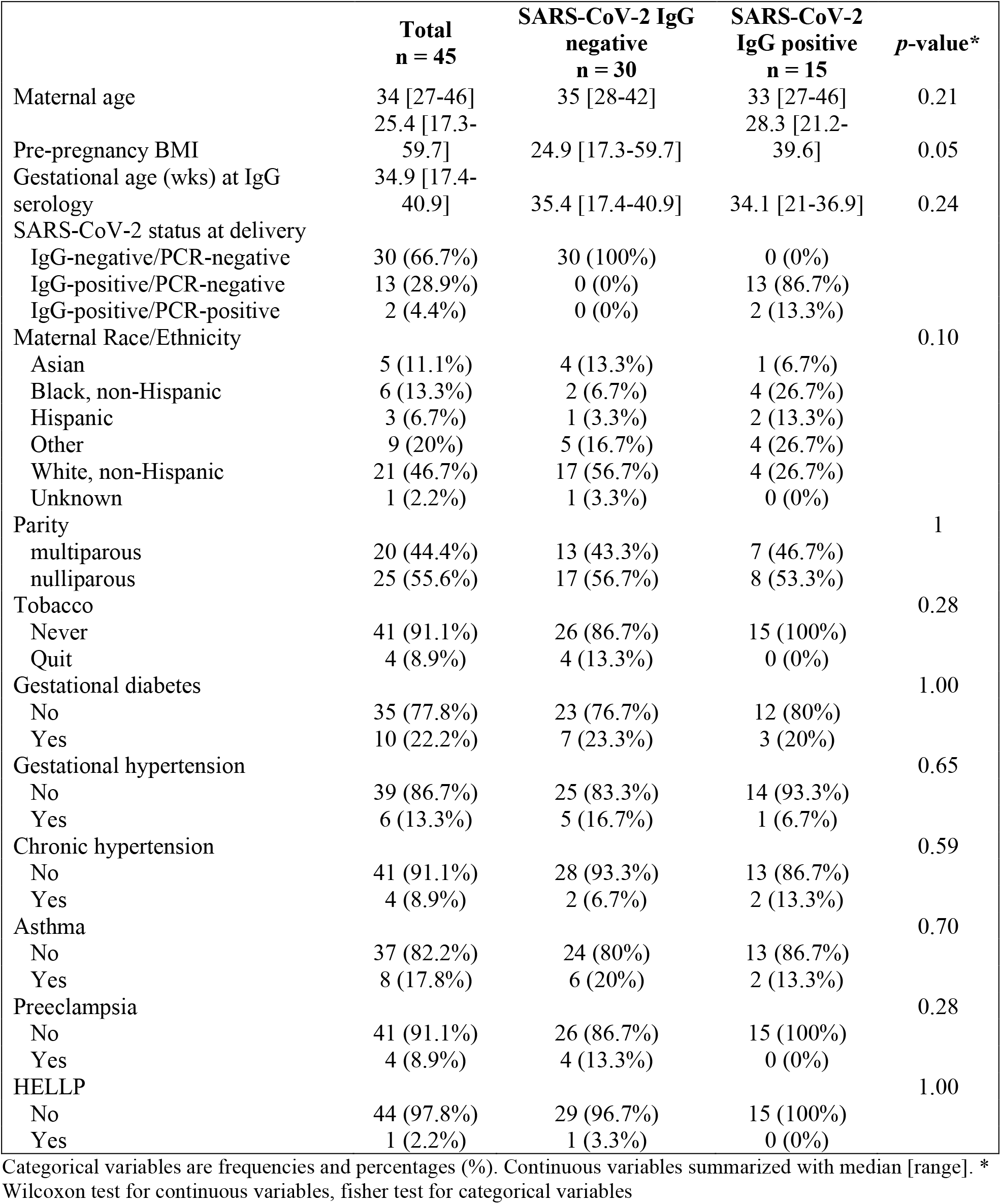
Maternal characteristics stratified by SARS-CoV-2 IgG status

The characteristics of the newborns are shown in **Table 2**. All were live births, 82.2% (n=37) delivered at term, and 17.8 delivered preterm. We did not detect differences in gestational age between study groups; median gestational age at delivery was 38.9 and 39 weeks for newborns born to IgG-negative and IgG-positive participants, respectively (*p*=0.87). The distribution of newborn sex was slightly different; 66.7% of newborns in the seropositive group were female and 33.3% were male, while in the seronegative group 33.3% were female and 66.7% were male (*p*=0.06). No differences were noted in birthweight, delivery mode, intrauterine growth restriction, APGAR scores, or NICU admission rates.

**Table 2.**
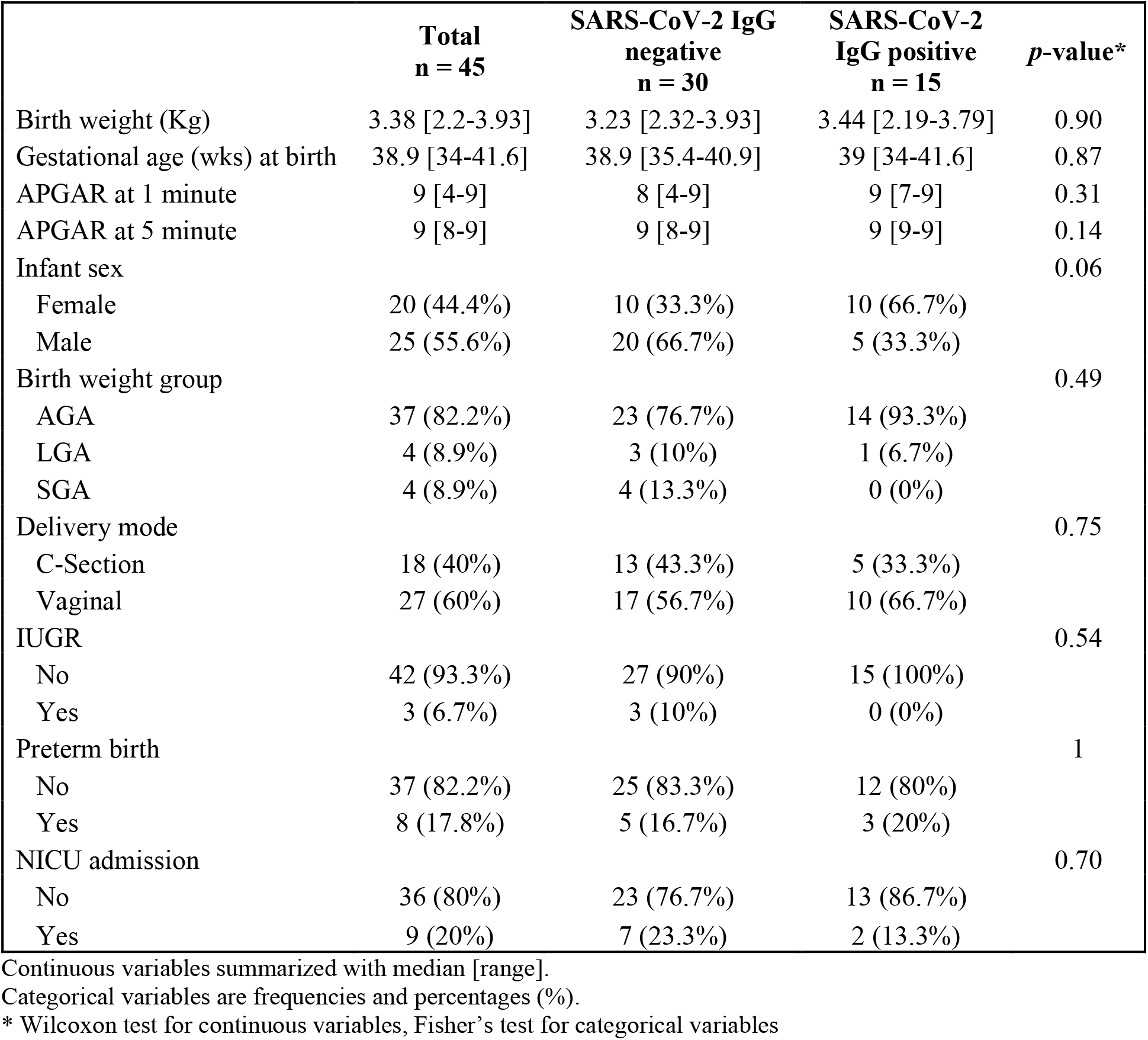
Newborn characteristics stratified by maternal SARS-CoV-2 IgG status

The pathology examination showed that placental weight was comparable between IgG seropositive and seronegative participants. Similarly, study groups were not different in other histopathology findings including chronic villitis, deciduitis, acute chorioamnionitis, intervillitis, intervillous thrombosis, fetal vascular thrombosis, decidual arteriopathy, fibrin presence, or chorangiosis (**Table 3**).

**Table 3.** Histopathology characteristics stratified by maternal SARS-CoV-2 IgG status

|  | **Total**<br>**n = 45** | **SARS-CoV-2 IgG**<br>**negative**<br>**n = 30** | **SARS-CoV-2**<br>**IgG positive**<br>**n = 15** | ***p*-value\*** |
| --- | --- | --- | --- | --- |
| Placental weight (g) | 501 [304-644] | 476 [304-644] | 504 [317-568] | 0.92 |
| Chronic villitis | 2 (4.4%) | 1 (3.3%) | 1 (6.7%) | 1 |
| Deciduitis | 2 (4.4%) | 2 (6.7%) | 0 (0%) | 0.55 |
| Acute chorioamnionitis | 10 (22.2%) | 6 (20%) | 4 (26.7%) | 0.71 |
| Intervillitis | 1 (2.2%) | 0 (0%) | 1 (6.7%) | 0.33 |
| Intervillous thrombosis | 2 (4.4%) | 2 (6.7%) | 0 (0%) | 0.55 |
| Fetal vascular thrombosis | 1 (2.2%) | 1 (3.3%) | 0 (0%) | 1.00 |
| Decidual arteriopathy | 5 (11.1%) | 5 (16.7%) | 0 (0%) | 0.15 |
| Fibrin | 32 (71.1%) | 22 (73.3%) | 10 (66.7%) | 0.73 |
| Chorangiogenesis | 2 (4.4%) | 2 (6.7%) | 0 (0%) | 0.55 |
| Any inflammatory lesion<sup>#</sup> | 14 (31.3%) | 7 (23.3%) | 7 (46.7%) | 0.17 |

Continuous variables summarized with median [range]. Categorical variables are frequencies and percentages (%).
\* Wilcoxon test for continuous variables, Fisher's exact test for categorical variables.

### 3.2 Placenta gene expression analysis

After preprocessing, quality control, the gene expression dataset consisted of 44 samples (IgG negative n=29; IgG positive n=15) and 48 genes. Nine genes were consistently below the background level in both study groups and were excluded from differential expression analyses. These included the nucleocapsid and the envelope viral SARS-CoV-2 genes, SARS-CoV-2 cell-entry genes (*TMPRSS2, ACE2*), and genes involved in the immune and stress response (*IL17A, IL23A, IFNL3, IFNA1* and *OPRM1*). Summary statistics for the expression of the 39 detected genes are shown in **Supplementary Table 2**. We used principal components analyses to identify effects of biologic and technical covariates in the overall placental gene expression patterns, which did not reveal obvious clustering by SARS-CoV-2 IgG serology or nasal swab PCR status at delivery (**Supplementary Fig. 1**).

Next, we performed differential expression analyses by SARS-CoV-2 IgG status adjusted for covariates and cell-type proxies (**Figure 1**). In the analyses adjusted for covariates only, three genes were significantly associated with SARS-CoV-2 IgG antibody status (**Figure 1A, Supplementary Table 3**). The trophoblast cell-markers *PSG3* and *CGB3* were downregulated (log2 fold-change [log2FC] = - 0.9, *p*=0.002 and log2FC=-0.99, *p*=0.05, respectively). *PSG3* and *CGB3* placental expression levels were highly correlated (rho=0.6, *p*=6.1×10^−5^). The chemokine *CXCL10* was over-expressed (log2FC = 1.08, *p*=0.02) in the IgG positive group compared to the IgG negative group. Since placental tissues are heterogenous mixtures of cells, we repeated the differential expression analyses adjusting for covariates and expression cell-type proxies including the average expression *PSG3* and *CGB3* as syncytiotrophoblast markers. In this analysis, SARS-CoV-2 IgG positivity was associated with increased expression of three genes *CXCL10, TLR3* and *DDX58* (**Figure 1B, Supplementary Table 4**). The differences in placental gene expression between the SARS-CoV-2 IgG positive and negative groups ranged between log2 FC of 1.19 for *CXCL10* and 0.30 for *DDX58* (**Figure 2**). Sensitivity analyses excluding samples from the two participants who were SARS-CoV-2 IgG positive and PCR positive at delivery showed similar results (**Supplemental Fig. 2**).

**Figure 1.**
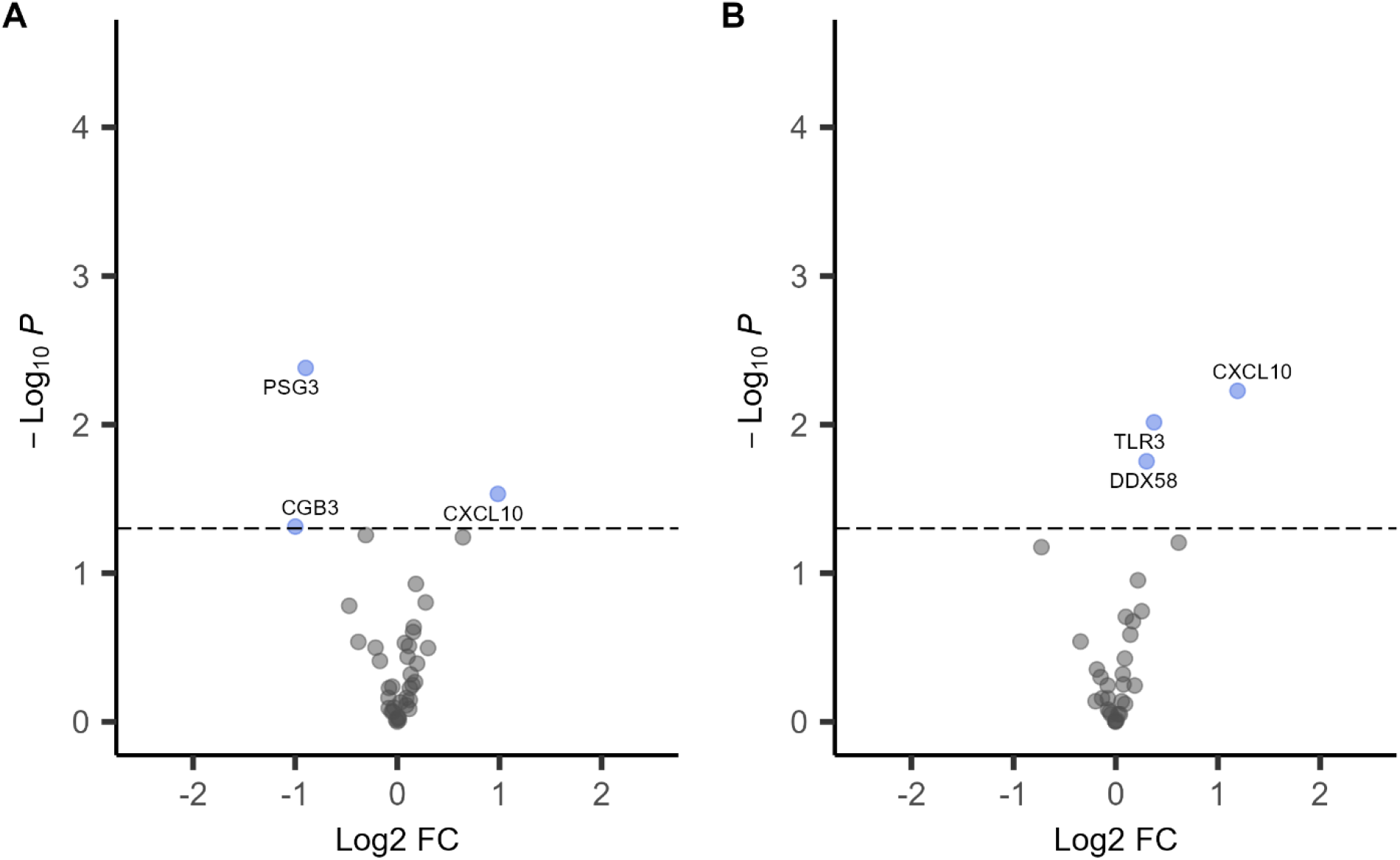
Volcano plots of differential gene expression analyses in placentas from SARS-CoV-2 IgG positive (n=15) versus IgG negative (n=29) participants. A. Linear models adjusted for infant sex, birthweight, gestational age at birth and maternal age. B. Linear models adjusted for infant sex, birthweight, gestational age at birth and maternal age and cell-type gene expression proxies. The x-axis is the log2 fold change (FC) and y-axis is the –log10 *P*-value, the highlighted in blue are genes with *p*<0.05. The horizontal dashed line corresponds to *p*=0.05.

**Figure 2.**
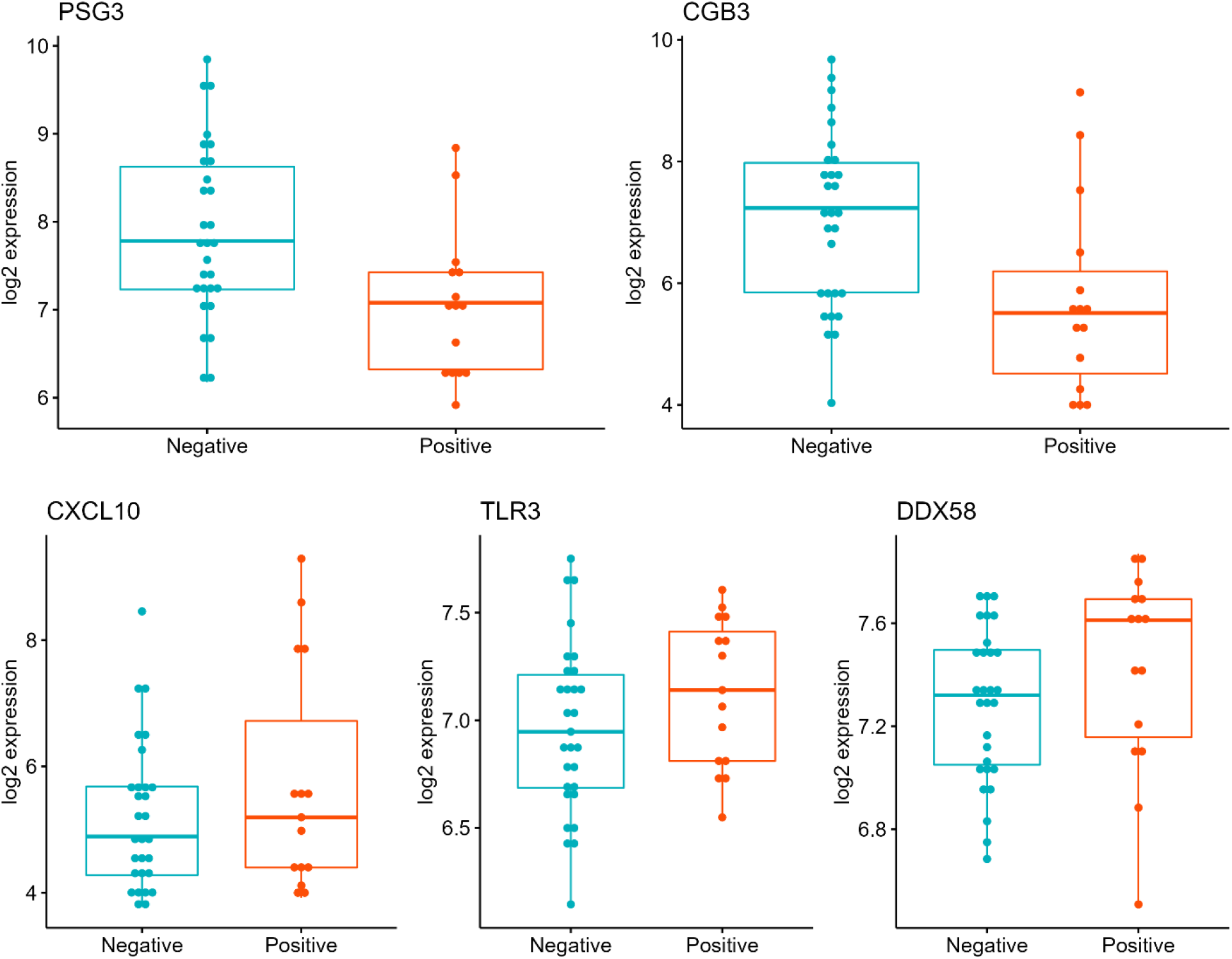
Box plots of gene expression in placentas delivered by SARS-CoV-2 IgG positive versus IgG negative participants. Top panel. *PSG3* and *CGB3* trophoblast genes. Bottom panel *CXCL10, TLR3* and *DDX58* immune genes. The y-axis is the log2 fold change (FC) and x-axis is SARS-CoV-2 IgG antibody group.

## 3. Discussion

The effects of SARS-CoV-2 infection on the placenta are not yet well-characterized, particularly in cases of infection during pregnancy without active infection at delivery (i.e., SARS-CoV-2 IgG positive and negative PCR at the delivery admission). In this report, we investigated histopathology and molecular gene expression changes in placentas from 15 mother-infant pairs exposed to SARS-CoV-2 during pregnancy and 30 unexposed controls, part of the Generation C study in NYC. We report differences in the expression of trophoblast specific and immune genes in placentas between IgG positive and negative mothers.

All the placentas in this study underwent medical pathology review, and we found a range of histopathologic findings. However, none of the findings differed significantly with respect to SARS-CoV- 2 IgG serology status, which is consistent with some studies on Covid-19 and placenta pathology [39, 51]. A recent systematic review and pooled analysis of case-control reports found increased risk of fetal vascular malperfusion, chronic inflammatory pathology, perivillous fibrin, and intervillous thrombosis [41], yet most of the reviewed cases in that analyses were RT-PCR positive at delivery. Our results are inconsistent with these findings, possibly due to limited sample size or because our study population consisted mostly of individuals who were infected with the virus during pregnancy without active infection at delivery.

We detected significant associations between plasma SARS-CoV-2 IgG positivity and lower placental expression of the *PSG3* and *CGB3*. These placenta-specific genes locate to 19q13 and are highly expressed by trophoblast cells. *PSG3* is part of the family of human pregnancy-specific glycoproteins (PSG) that are released to the maternal circulation during pregnancy and are reported to have immunoregulatory and angiogenic functions [52]. *CGB3* encodes the beta 3 subunit of the chorionic gonadotropin (hCG), a glycoprotein hormone essential for pregnancy maintenance, that has also been involved in angiogenesis and maternal immunotolerance [53, 54]. Some studies have reported associations between adverse pregnancy outcomes like preeclampsia and circulating levels of PSGs and hCG [52, 53, 55, 56]. Importantly, *PSG3* and *CGB3* are involved in trophoblast syncytialization; alterations in expression of these genes have been described in other viral infections [57, 58]. However, to our knowledge, there are no studies linking past maternal SARS-CoV-2 infection to altered placental *PSG3* or *CGB3* expression or trophoblast differentiation.

We also observed increased expression of *CXCL10* in placentas delivered from participants exposed to SARS-CoV-2 infection during pregnancy. *CXCL10* (C-X-C motif chemokine ligand 10) is a pro-inflammatory chemokine secreted in response to interferon gamma (IFNγ) involved in the stimulation of monocytes, natural killer, and T-cells. Previous investigations have also reported *CXCL10* gene expression upregulation in bronchoalveolar lavages [57], nasopharyngeal swabs [58] and male placentas exposed to maternal SARS-CoV-2 infection [43]. Importantly, *CXCL10* may also be a key regulator of the “cytokine storm” in response to SARS-CoV-2 infection and circulating levels of CXCL10 are reported to be positively associated with disease severity [59-61]. Two other immune genes, *TLR3* and *DDX58*, were overexpressed in placentas delivered from SARS-CoV-2 IgG positive participants. *TLR3* is a member of the toll-like receptor (TLR) family and *DDX58* encodes a protein containing RNA helicase-DEAD box motifs (also known as RIG-I). These genes are involved in recognizing double-stranded RNA (dsRNA) released during viral replication and activation of the innate immune response [62, 63]. To date, placental expression of these genes has not been linked to SARS-CoV-2 infection. However, peripheral blood *TLR3* gene expression is reduced in patients with severe COVID-19 compared to those with mild forms of the disease [64] and *DDX58* expression in human lung cells has been implicated in the initial response against SARS-CoV-2 infection [65, 66].

Strengths of the Generation C study include a demographically diverse population of pregnant participants recruited in NYC after the start of the SARS-CoV-2 pandemic. To assess SARS-CoV-2 IgG levels, we used a highly sensitive (95%) and specific (100%) serological assay. Also, we explored the effects of SARS-CoV-2 IgG seropositivity on placental histopathology and gene expression simultaneously. We acknowledge that our study is not without limitations. The sample size is limited, and the number of placentas delivered from SARS-CoV-2 IgG positive participants is small. We do not have information on the precise timing of SARS-CoV-2 infection, disease severity, or newborn SARS-CoV-2 IgG levels. Given the exploratory nature of this study, we only measured expression of a small number of genes, and we did not account for multiple testing. Larger studies are needed to investigate the effects of SARS-CoV-2 infection on the whole placental transcriptome. In summary, we found evidence of an association between SARS-CoV-2 infection during pregnancy and placental expression of trophoblast and immune related genes which could potentially contribute to long-term health effects in the offspring. Future research should confirm the observed associations and assess potential long-term implications.

## Data Availability

All data produced in the present study are available upon reasonable request to the authors

## Funding

The Generation C cohort was established through funding from the US Centers for Disease Control and Prevention (CDC), who also provided technical assistance related to analysis and interpretation of data and writing the report (contract 75D30120C08186). CL is funded through NIH-NICHD R00HD097286. The findings and conclusions in this report are those of the authors and do not necessarily represent the position of the funding agencies. The findings and conclusions in this report are those of the authors and do not necessarily represent the official position of the CDC.

## Declaration of competing interest

None.

## Acknowledgements

We would like to thank all participants of the Generation C study for their cooperation and contribution to the research field. We would like to thank the members of the Krammer Serology Core Study group and the Mount Sinai Biorepository and Pathology Core, and especially Maryann Huie, Frances Avila, Ariane Benedetto, Anastasiya Dzhun.

## Notes

### Competing Interest Statement

The authors have declared no competing interest.

### Author Declarations

Institutional Review Board at the Icahn School of Medicine at Mount Sinai (protocol IRB-20-03352).

