## Appendix for "Gestational SARS-CoV-2 infection is associated with placental expression of immune and trophoblast genes"

**Supplementary Table 1.** Genes profiled in the placenta using a custom-designed NanoString codeset

| Category | # of genes | Gene Symbols |
|---|---|---|
| Immune/inflammatory response genes | 25 | *CCL2*, *CD14*, *CXCL10*, *CXCL8*, *DDX58*, *EGR1*, *ELK1*, *HSPD1*, *IFNA1*, *IFNL3*, *IL17A*, *IL1B*, *IL23A*, *IL6*, *IRAK2*, *IRF3*, *NFKB2*, *NFKBIL1*, *NKG7*, *TIRAP*, *TLR3*, *TLR7*, *TNF*, *TOLLIP* |
| Stress response genes | 8 | *DYRK1A*, *FKBP5*, *HSD11B2*, *MAOA*, *NR3C1*, *NR3C2*, *OGT*, *OPRM1* |
| Cell type marker genes | 7 | *CD163*, *CD68*, *CGB3*, *PECAM1*, *PEG10*, *PEG3*, *PSG3* |
| Covid-19 host response genes | 6 | *ACE2*, *HOPX*, *DHX58*, *FURIN*, *HAS2*, *TMPRSS2* |
| Viral genes | 2 | SARS-CoV-2_E, SARS-CoV-2_N |
| Housekeeping genes* | 2 | *RPL19*, *RPLP0* |

* used for normalization

**Supplementary Table 2**. Descriptive statistics of placental gene expression.

| Gene | mean | SD | median | min | max | N above LOD[1] | % above LOD |
|---|---|---|---|---|---|---|---|
| *PEG10* | 13.20 | 0.46 | 13.19 | 12.19 | 14.51 | 48 | 1 |
| *PECAM1* | 12.49 | 0.39 | 12.50 | 11.56 | 13.15 | 48 | 1 |
| *PEG3* | 11.72 | 0.27 | 11.74 | 11.13 | 12.30 | 48 | 1 |
| *CD14* | 10.86 | 0.68 | 10.89 | 8.96 | 12.42 | 48 | 1 |
| *HSPD1* | 10.42 | 0.25 | 10.38 | 9.95 | 11.10 | 48 | 1 |
| *NR3C1* | 10.34 | 0.44 | 10.33 | 9.42 | 11.73 | 48 | 1 |
| *CD163* | 9.82 | 0.62 | 9.90 | 8.24 | 11.59 | 48 | 1 |
| *OGT* | 9.52 | 0.34 | 9.53 | 8.94 | 10.35 | 48 | 1 |
| *CD68* | 9.23 | 0.65 | 9.29 | 7.25 | 10.32 | 48 | 1 |
| *FURIN* | 9.15 | 0.44 | 9.06 | 8.51 | 10.07 | 48 | 1 |
| *DYRK1A* | 8.85 | 0.21 | 8.87 | 8.39 | 9.27 | 48 | 1 |
| *CCL2* | 8.50 | 0.94 | 8.75 | 6.07 | 10.58 | 48 | 1 |
| *FKBP5* | 8.85 | 0.87 | 8.59 | 7.34 | 11.21 | 48 | 1 |
| *EGR1* | 8.59 | 1.10 | 8.56 | 5.86 | 11.22 | 47 | 0.98 |
| *NFKB2* | 8.57 | 0.40 | 8.52 | 7.89 | 9.71 | 48 | 1 |
| *TOLLIP* | 8.29 | 0.25 | 8.29 | 7.73 | 9.12 | 48 | 1 |
| *HOPX* | 8.00 | 0.58 | 8.03 | 6.75 | 9.11 | 48 | 1 |
| *MAOA* | 8.02 | 0.62 | 8.00 | 6.90 | 9.54 | 48 | 1 |
| *IRF3* | 7.81 | 0.40 | 7.74 | 7.31 | 9.97 | 48 | 1 |
| *PSG3* | 7.60 | 1.00 | 7.41 | 5.92 | 9.85 | 48 | 1 |
| *DDX58* | 7.33 | 0.33 | 7.35 | 6.51 | 7.87 | 47 | 0.98 |
| *HSD11B2* | 7.41 | 0.89 | 7.21 | 5.89 | 9.11 | 47 | 0.98 |
| *TLR3* | 7.02 | 0.38 | 7.03 | 6.15 | 7.75 | 47 | 0.98 |
| *CXCL8* | 7.20 | 1.53 | 6.95 | 4.98 | 10.71 | 47 | 0.98 |
| *IL6* | 6.60 | 0.66 | 6.63 | 5.53 | 8.14 | 47 | 0.98 |
| *CGB3* | 6.67 | 1.53 | 6.58 | 3.31 | 9.68 | 47 | 0.98 |
| *TIRAP* | 6.53 | 0.38 | 6.57 | 5.88 | 7.50 | 47 | 0.98 |
| *NKG7* | 6.25 | 0.98 | 6.41 | 3.27 | 8.26 | 46 | 0.96 |
| *NFKBIL1* | 6.26 | 0.60 | 6.30 | 3.25 | 7.53 | 46 | 0.96 |
| *ELK1* | 6.34 | 0.44 | 6.28 | 5.47 | 7.79 | 47 | 0.98 |
| *IL1B* | 5.88 | 1.39 | 5.99 | 2.77 | 8.78 | 44 | 0.92 |
| *NR3C2* | 5.87 | 0.75 | 5.98 | 3.25 | 6.83 | 45 | 0.94 |
| *HAS2* | 5.78 | 0.96 | 5.75 | 3.25 | 7.67 | 44 | 0.92 |
| *DHX58* | 5.54 | 0.66 | 5.70 | 3.25 | 6.46 | 44 | 0.92 |
| *CXCL10* | 5.31 | 1.53 | 5.18 | 2.77 | 9.29 | 38 | 0.79 |
| *TLR7* | 4.88 | 0.85 | 5.09 | 2.77 | 6.41 | 39 | 0.81 |
| *IRAK2* | 4.55 | 0.66 | 4.62 | 3.25 | 5.97 | 36 | 0.75 |
| *IL10* | 4.49 | 1.12 | 4.47 | 2.27 | 7.56 | 31 | 0.65 |
| *TNF* | 4.62 | 1.19 | 4.34 | 2.77 | 7.15 | 33 | 0.69 |
| *IFNL3* | 3.75 | 0.66 | 3.63 | 2.27 | 6.24 | 12 | 0.25 |
| *IFNA1* | 3.82 | 0.81 | 3.57 | 2.77 | 7.21 | 8 | 0.17 |
| *TMPRSS2* | 3.71 | 0.72 | 3.55 | 2.45 | 6.72 | 5 | 0.10 |
| *ACE2* | 3.57 | 0.56 | 3.54 | 2.15 | 5.79 | 4 | 0.08 |
| *IL23A* | 3.61 | 0.57 | 3.53 | 2.27 | 5.87 | 3 | 0.06 |
| *OPRM1* | 3.66 | 0.67 | 3.53 | 2.27 | 6.39 | 6 | 0.13 |
| *IL17A* | 3.51 | 0.35 | 3.51 | 2.27 | 4.16 | 0 | 0.00 |

**LOD:** limit of background expression detection.

**Supplementary Table 3.** Differential expression analyses results─ analyses adjusted for covariates

| Gene | Log FC | *p*-value | Adjusted *p*-value |
|---|---|---|---|
| *PSG3* | -0.90 | 0.004 | 0.16 |
| *CXCL10* | 0.98 | 0.029 | 0.45 |
| *CGB3* | -1.00 | 0.05 | 0.45 |
| *PEG10* | -0.31 | 0.06 | 0.45 |
| *NKG7* | 0.64 | 0.06 | 0.45 |
| *DDX58* | 0.18 | 0.12 | 0.77 |
| *IFNL3* | 0.28 | 0.16 | 0.81 |
| *HSD11B2* | -0.47 | 0.17 | 0.81 |
| *PECAM1* | 0.16 | 0.23 | 0.83 |
| *TLR3* | 0.15 | 0.25 | 0.83 |
| *EGR1* | -0.38 | 0.29 | 0.83 |
| *DYRK1A* | 0.07 | 0.29 | 0.83 |
| *OGT* | 0.11 | 0.31 | 0.83 |
| *ELK1* | -0.21 | 0.32 | 0.83 |
| *FKBP5* | 0.30 | 0.32 | 0.83 |
| *NFKB2* | 0.10 | 0.37 | 0.88 |
| *HOPX* | -0.17 | 0.39 | 0.88 |
| *TLR7* | 0.19 | 0.41 | 0.88 |
| *IRAK2* | 0.13 | 0.48 | 0.96 |
| *DHX58* | 0.17 | 0.54 | 0.96 |
| *TIRAP* | 0.15 | 0.57 | 0.96 |
| *PEG3* | -0.05 | 0.58 | 0.96 |
| *FURIN* | -0.08 | 0.59 | 0.96 |
| *NFKBIL1* | 0.12 | 0.59 | 0.96 |
| *CD68* | -0.09 | 0.69 | 0.98 |
| *CD163* | 0.09 | 0.69 | 0.98 |
| *HAS2* | 0.12 | 0.71 | 0.98 |
| *TOLLIP* | 0.03 | 0.74 | 0.98 |
| *CCL2* | 0.09 | 0.77 | 0.98 |
| *IRF3* | -0.03 | 0.80 | 0.98 |
| *MAOA* | -0.08 | 0.81 | 0.98 |
| *CXCL8* | 0.12 | 0.82 | 0.98 |
| *NR3C2* | -0.06 | 0.85 | 0.98 |
| *CD14* | -0.04 | 0.85 | 0.98 |
| *NR3C1* | 0.01 | 0.93 | 1.00 |
| *HSPD1* | 0.01 | 0.95 | 1.00 |
| *TNF* | -0.01 | 0.97 | 1.00 |
| *IL1B* | 0.01 | 0.97 | 1.00 |
| *IL6* | 0.00 | 1.00 | 1.00 |

*Models adjusted for infant sex, birthweight, gestational age at birth and maternal ag. Log FC: log2 fold change, adjusted p-value: false discovery rate adjusted p-value.

**Supplementary Table 4.** Differential expression analyses results--analyses adjusted for covariates and cell-type gene expression proxies*

| Gene | Log FC | *p*-value | Adjusted *p*-value |
|---|---|---|---|
| *CXCL10* | 1.19 | 0.01 | 0.15 |
| *TLR3* | 0.37 | 0.01 | 0.15 |
| *DDX58* | 0.30 | 0.02 | 0.19 |
| *NKG7* | 0.62 | 0.06 | 0.43 |
| *EGR1* | -0.73 | 0.07 | 0.43 |
| *FURIN* | 0.22 | 0.11 | 0.60 |
| *IFNL3* | 0.25 | 0.18 | 0.75 |
| *DYRK1A* | 0.10 | 0.20 | 0.75 |
| *IRF3* | 0.17 | 0.21 | 0.75 |
| *OGT* | 0.14 | 0.26 | 0.83 |
| *IL6* | -0.34 | 0.29 | 0.84 |
| *TOLLIP* | 0.09 | 0.38 | 1.00 |
| *ELK1* | -0.18 | 0.44 | 1.00 |
| *HSPD1* | 0.07 | 0.48 | 1.00 |
| *TLR7* | -0.15 | 0.50 | 1.00 |
| *NFKB2* | 0.08 | 0.56 | 1.00 |
| *CD14* | -0.08 | 0.57 | 1.00 |
| *FKBP5* | 0.19 | 0.57 | 1.00 |
| *MAOA* | -0.13 | 0.69 | 1.00 |
| *IRAK2* | -0.08 | 0.70 | 1.00 |
| *CXCL8* | -0.20 | 0.73 | 1.00 |
| *HOPX* | 0.06 | 0.73 | 1.00 |
| *HSD11B2* | 0.09 | 0.76 | 1.00 |
| *NR3C2* | -0.07 | 0.83 | 1.00 |
| *CCL2* | -0.06 | 0.86 | 1.00 |
| *NR3C1* | 0.02 | 0.89 | 1.00 |
| *TIRAP* | 0.04 | 0.89 | 1.00 |
| *TNF* | -0.05 | 0.89 | 1.00 |
| *NFKBIL1* | 0.01 | 0.97 | 1.00 |
| *DHX58* | 0.00 | 0.99 | 1.00 |
| *HAS2* | 0.00 | 0.99 | 1.00 |
| *IL1B* | 0.00 | 1.00 | 1.00 |

*Models adjusted for infant sex, birthweight, gestational age at birth, maternal age and cell type gene marker (PEG10 & PEG3 for cytotrophoblasts, CGB3 & PSG3 for syncytiotrophoblasts, CD68 & CD163 for macrophages, PECAM1 for endothelial cells). Log FC: log2 fold change, adjusted p-value: false discovery rate adjusted p-value.

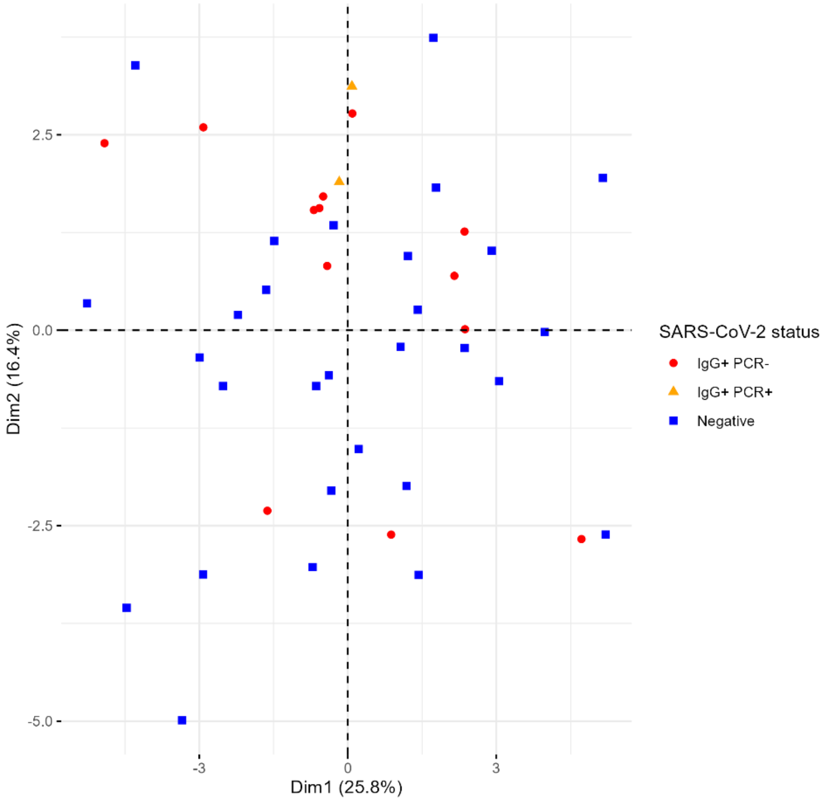


**Supplementary figure 1**. Principal component analyses of placenta gene expression data by plasma SARS-CoV-2 IgG antibody (median gestational age 35 weeks) and nasal swab SARS-CoV-2 PCR (at delivery).

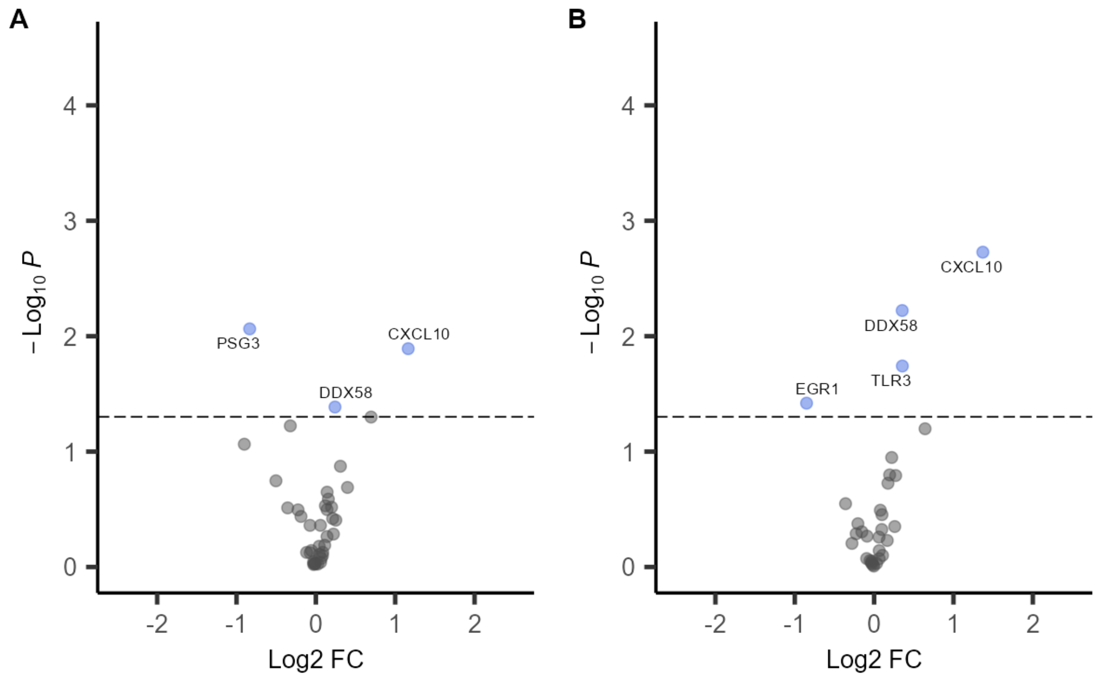


**Supplementary figure 2.** Volcano plots of differential gene expression analyses of placentas delivered by SARS-CoV-2 IgG positive (n=13) versus IgG negative (n=29) participants data from sensitivity analyses after excluding the 2 placenta samples that were PCR positive at delivery. A. Linear models adjusted for infant sex, birthweight, gestational age at birth and maternal age. B. Linear models adjusted for infant sex, birthweight, gestational age at birth and maternal age and cell-type gene expression proxies. The x-axis is the log2 fold change (FC) and y-axis is the –log10 P-value, the highlighted in blue are genes with p<0.05. The horizontal dashed line corresponds to p=0.05.
